# Developing a GraphRAG-enabled local-LLM for Gestational Diabetes Mellitus

**DOI:** 10.1101/2025.04.28.25326568

**Authors:** Edmund Evangelista, Fathima Ruba, Syed M. Salman Bukhari, Amril Nazir, Ravi S. Sharma

## Abstract

This paper re-imagines a world of abundance in the treatment of chronic diseases such as Tpe 2 Diabetes. It asks: what if preventive and diagnostic remedies were widely made available across the world, informed by the latest medical research? As Proof-of-Concept of a proposed solution, the paper describes the development and validation of a local Large Language Models (local-LLMs) based on Graph-based Retrieval-Augmented Generation (GraphRAG) for managing Gestational Diabetes Mellitus (GDM). The research thus seeks new insights into optimizing GDM treatment through a knowledge graph architecture, contributing to a deeper understanding of how artificial intelligence can extend medical expertise to underserved populations globally. The study employs an agile, prototyping approach utilizing GraphRAG to enhance knowledge graphs by integrating retrieval-based and generative artificial intelligence techniques. Training data was from academic papers published between January 2000 and May 2024 using the Semantic Scholar API and analyzed by mapping complex associations within GDM management to create a comprehensive knowledge graph architecture. It is categorically stated that, since the primary research objective was to establish the feasibility of a GraphRAG local-LLM PoC, no human subjects nor actual patient datasets were used. Empirical results indicate that the GraphRAG-based Proof of Concept outperforms open-source LLMs such as ChatGPT, Claude, and BioMistral across key evaluation metrics. Specifically, GraphRAG achieves superior accuracy with BLEU scores of 0.99, Jaccard similarity of 0.98, and BERT scores of 0.98, offering significant implications for personalized medical insights that enhance diagnostic accuracy and treatment efficacy. This research offers a novel perspective on applying GraphRAG-enabled LLM technologies to GDM management, providing valuable insights that extend current understanding of AI applications in healthcare. The study’s findings contribute to advancing the feasibility of GenAI for proactive GDM treatment and extending medical expertise to underserved populations globally.

## 1. Introduction

The growing use of electronic medical records (EMR) linking diverse patient characteristics and prescription choices with positive treatment outcomes in large-scale use cases has revealed the potential to benchmark the effectiveness of medical options and inform optimal choices for personalized medicine. For example, Sharma et al. (2020) outlined an approach for delivering personalized healthcare as a means of effectively utilizing scarce medical resources in underserved regions and populations, reinforcing the value of AI-driven systems in such settings. While machine learning and optimization techniques have generated individualized treatment recommendations for improving outcomes, “these works focused on making broad [largely drug class level] treatment recommendations independently of specific drug and dose considerations … [whereas] guidelines and landmark trials highlight important drug– and dose-dependent variations in treatment efficacy, safety, and risk profiles” (Nambiar **et al.** 2024). In short, personalized medicine should account for contextual variations in seeking more effective treatments, costs, and outcomes. This paper presents an approach to personalized medicine using a generative AI approach to treat a chronic condition afflicting young mothers and their children with increasing alacrity. Such a need is particularly acute in the socio-economically disadvantaged regions of the world.

Gestational Diabetes Mellitus (GDM) is a significant global health concern affecting a substantial number of pregnancies (Kjos and Buchanan, 1999). Defined as glucose metabolism imbalance first detected during pregnancy, the International Association of Diabetes in Pregnancy Study Group (IADPSG) reports that “GDM is not only related to perinatal morbidity but also to an increased risk of diabetes and cardiovascular disease in the mother in later life, and childhood obesity in the offspring” (Metzger et al. 2010). The pooled global prevalence was 14% in 2021 with the highest occurrence in the Middle East – North Africa (27.6%), Southeast Asia (20.8%), and among high-income countries (14.2%) (Akinyemi, 2024). There is considerable agreement among medical practitioners that the development of GDM could be influenced by various risk factors, including maternal age, obesity, family history of diabetes, previous occurrences of GDM, and certain ethnic backgrounds (Bashir and Bajwa, 2021; Ejaz et al. 2023). This is depicted in Figure 1 (data sources: (Kjos and Buchanan, 1999; Macaulay et al. 2014; Stewart-Field, 2023; Kurt et al. 2023)) as the medical characterization of GDM comprising factors such as diagnosis, risks, prediction, management, complication, and post-partum care.

**Figure 1:**
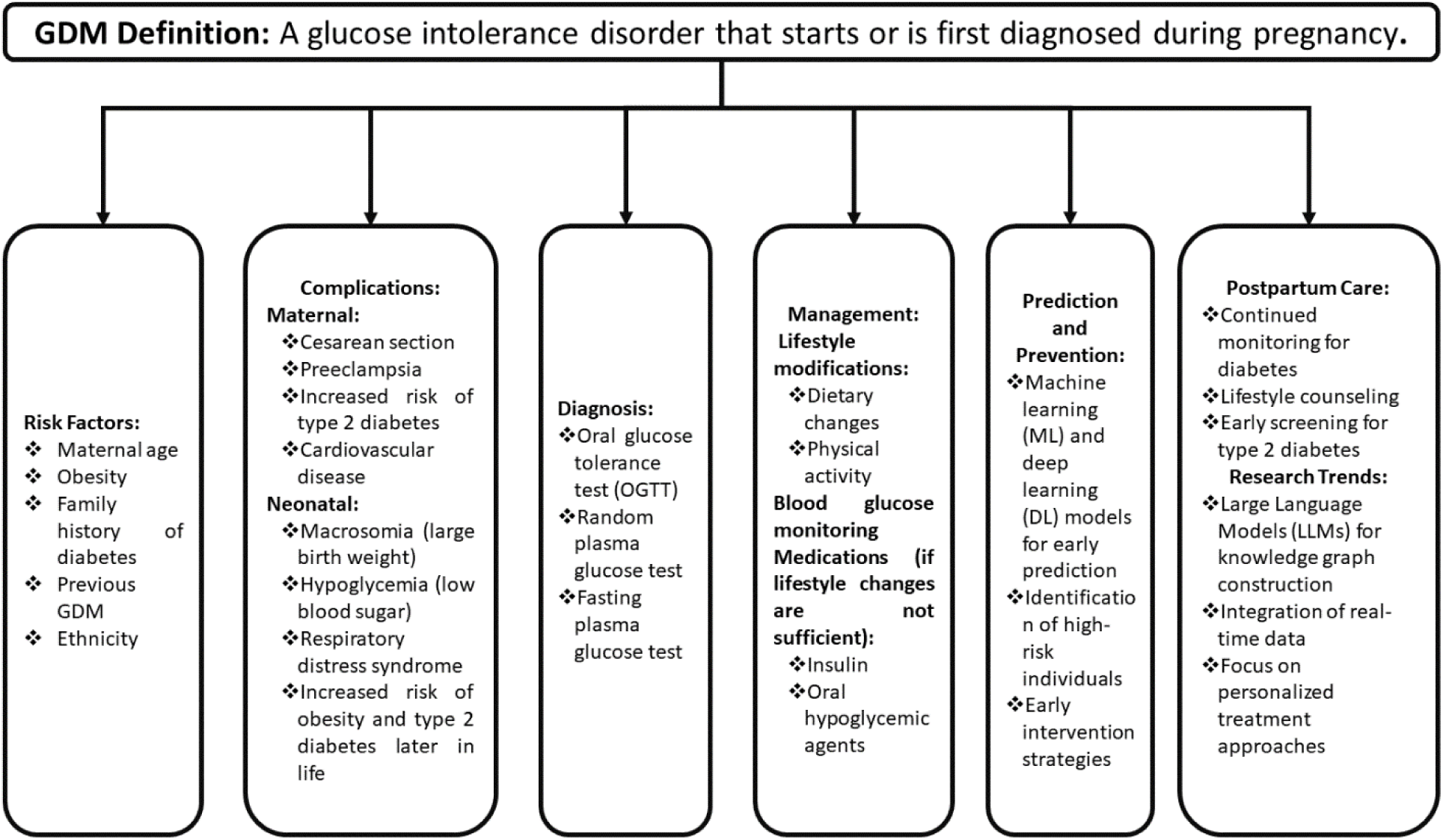
Medical Characterization of Gestational Diabetes Mellitus (GDM).

Also of concern to the WHO is that GDM leads to various complications for both mothers and children, such as increased risks of cesarean delivery, preeclampsia, and type 2 diabetes (T2D) for mothers. Children are at higher risk of macrosomia, hypoglycemia, respiratory distress syndrome, and an increased likelihood of developing obesity and T2D later in life (Jani et al. 2023). Long-term health risks include elevated chances of developing T2D and cardiovascular diseases for both mother and child (Alum et al. 2024). In the Global South and other developing countries (Macaulay et al. 2014; Hinneh et al. 2022), GDM presents significant challenges due to:

1. Higher prevalence rates in certain regions, particularly South Asia and the Middle East.
2. Limited healthcare resources for screening, diagnosis, and management.
3. Genetic factors in certain ethnic groups that increase GDM risk.
4. Rapid urbanization and lifestyle changes leading to increased obesity rates.
5. Potential underdiagnosis due to lack of routine screening.

Effective GDM treatment requires multiple diagnostic tests, including oral glucose tolerance tests (OGTT), random plasma glucose tests, and fasting plasma glucose tests. Treatment options encompass regular blood glucose monitoring, dietary modifications, lifestyle changes, and, when necessary, pharmacological interventions such as insulin or oral hypoglycemic agents (StewartField, 2023). Recent advancements in AI-driven tools, such as the AI Drug Mix and Dose Advisor (AIDA) developed for T2D (Nambiar et al. 2024), have shown potential in optimizing pharmacological interventions by customizing drug and dose recommendations to individual patient profiles. Similar approaches could be valuable in improving glycemic management in GDM cases, enhancing personalized care in postpartum treatment, drug discovery with therapy, and reducing long-term risks of developing chronic diseases in general^1^.

Given the technology trends and current research gaps, this paper reports a GenAI solution for GDM management in terms of early detection, access to evidence-based research findings, and personalized treatment options. Aside from bias and availability, current AI-based models mostly cannot integrate complex, multi-source medical data into actionable insights, leading to delays in diagnosis and resulting in suboptimal management of GDM. For the extraction and integration of intervention strategies from peer-reviewed medical research, we created a PoC of a knowledge graph (KG) that is powered by local Large Language Model (local LLM) to provide physicians with clinical support understand available treatment options.

Standalone local LLMs face limitations, such as producing hallucinations and confusion with complex, domain-specific queries (Pal et al. 2023). They underscore the need for the use or application of AI in context. To resolve these issues, we introduce Retrieval-Augmented Generation (RAG), which enhances the accuracy and relevance of the local LLM’s outputs by integrating external, contextual information (Miao et al. 2024; Dey et al. 2022). By efficiently combining diverse and complex data sources into a structured framework, our approach significantly improves predictive accuracy and offers valuable clinical support for personalized medical care. It is stated at the onset that since the primary research objective is to develop a PoC, any field validation of such a clinical support system using human subjects, live patient cases, and attending health practitioners is out of the intended scope of research.

This paper also investigates the boundaries of existing predictive models by utilizing generative tools in medical contexts, potentially transforming GDM treatment and enhancing public awareness through the strategic implementation of what is known as AI-generated K-Graphs (KG). This establishes a new benchmark for the monitoring and control of specialized and accurate healthcare interventions for GDM and possibly any other chronic diseases.

The remainder of this paper is organized as follows: This section provided an introduction to the research challenge that is to be addressed. The next section examines recent research on predicting and managing GDM, focusing on areas our KG RAG (or GraphRAG) approach aims to address. In Section 3, we detail our method of developing a PoC local LLM to generate and analyze KGs. This is followed by a discussion of our results which are benchmarked with widely adopted LLMs. We conclude in Section 5 by examining the implications of our findings for the future of GDM care and the broader field of AI-driven healthcare interventions.

## 2. Review of Background Research

There is little disagreement among health professionals that GDM presents significant challenges in prenatal healthcare, especially in settings with limited resources, where various factors further complicate the diagnosis and treatment of the disease (cf. Macaulay **et al.** 2014; Hinneh **et al.** 2022). Timely and accurate identification is essential for protecting the well-being of both the mother and the child. In areas with restricted availability of specialized services, challenges become more severe. Research in the area emphasizes the systemic problems that exacerbate these concerns in the Global South, such as a lack of standardized guidelines and limited access to essential screening and diagnostic equipment (Hinneh **et al.** 2022).

Additionally, there can be a shortage of healthcare professionals with expertise in GDM management, resulting in delays in identifying the condition and insufficient treatment. Economic limitations also have an important effect on the affordability of GDM care, while cultural beliefs and practices influence the extent to which patients adhere to treatment and engage in their healthcare (Kaewchandee and Vichitkunakorn, 2021; Sahu **et al.** 2021). A lack of knowledge and training on GDM among healthcare professionals and awareness among the general public leads to its underdiagnoses and mismanagement, which further affects the proper treatment of the condition (Watson **et al.** 2024). These issues highlight the necessity for improved clinical support for healthcare providers, seamless integration of GDM care into current healthcare systems, and the creation of interventions that are specifically designed to address the distinct requirements of these populations (Morampudi **et al.** 2017; Mpondo **et al.** 2015).

Recent advancements in Machine Learning (ML) and Deep Learning (DL) have clinically demonstrated the ability to improve the diagnosis and management of chronic diseases in recent years. These algorithms provide the capability to recognize individuals with a high risk of health-related issues at an early stage of pregnancy, customize treatment plans to their specific needs, and enhance the general well-being and health of both mothers and children. Several research efforts have developed ML algorithms that accurately predict GDM by accounting for demographic differences, such as a focus on Asian women (Kokori **et al.** 2024; Kumar **et al.** 2022). Kurt **et al. (**2023) proposed a clinical diagnostic system that uses decision trees and ensemble techniques specifically designed for the early diagnosis of GDM. This approach achieved a high level of sensitivity and specificity. However, these models fail to completely capture the complex connections that are inherent in GDM components.

Wu **et al. (**2021) addressed the importance of clinically interpretable ML methods and highlighted their predictive abilities in the monitoring and management of GDM. A similar study (Mennickent **et al.** 2023) on ML-based models for GDM prediction emphasized the need for high-quality datasets and thorough validation methods; it also identified major shortcomings in model interpretability and integration into clinical practice. The findings of Lu **et al. (**2023) criticized the incremental progress of integrating ML technology into digital health apps for controlling GDM, highlighting the urgent need for models that can be easily understood and interpreted by practitioners. The study by Pinto **et al. (**2023) examined the ability of XGBoost and explainable ML models to identify important risk factors for GDM. However, the absence of real-time intervention capabilities restricted the practical usefulness of their results. Cubillos **et al. (**2023) showed the capabilities of ML models in predicting GDM early on using maternal characteristics and blood biomarkers. However, it also noted that integrating these models into health data systems is currently limited.

Table I highlights some of the advancements and ongoing challenges in ML and DL models for predicting GDM. Generally, the table reveals the limitations ML models face in integrating various data sources and providing actionable, personalized insights. Many of these models lack interpretability, which makes them difficult to use in clinical settings, and they do not support real-time interventions, limiting their practical use. However, GenAI, especially through LLMs and their derivatives, has the potential to overcome these challenges by combining complex data and generating more accurate, context-relevant predictions. Local LLMs can improve treatment personalization and enhance GDM management by offering deeper insights into individual patient needs, promoting a more proactive and adaptable healthcare approach. This potential is already being realized in different healthcare sectors, where local LLMs and AI-driven solutions are being employed to address complex medical issues, as shown in recent studies (Nazi and Peng, 2024).

**Table I:** Representative Research DL/ML Models for Predicting GDM.

| Reference | Year | Model | Key Contributions (KCs) and Limitations (Limits.) |
|---|---|---|---|
| (Kokori *et al.* 2024; Kumar *et al.* 2022) | 2024 | Demographic-Specific ML Model | KCs.: Accurate predictions for specific demographics (Asian women).<br>Limits.: Limited integration into healthcare systems. |
| (Kurt *et al.* 2023) | 2023 | Decision Trees and Ensemble | KCs: High sensitivity and specificity.<br>Limits.: Fails to capture all GDM-related factors. |
| (J. Wu *et al.* 2024) | 2024 | Clinically Interpretable ML | KCs: Emphasized interpretable models for GDM.<br>Limits.: Limited real-time application. |
| (Mennickent *et al.* 2023) | 2022 | ML-Based Models | KCs: Importance of high-quality datasets.<br>Limits. : Lacks interpretability and integration. |

Advanced AI technologies such as Agentic AI and contextual AI using RAG have significantly transformed the processing, interpretation, and application of medical data. These innovations enable healthcare providers to efficiently analyze vast amounts of information, leading to earlier disease detection, more accurate diagnoses, and personalized treatment plans. For example, an AI model personalized for liver diseases has improved both diagnostic accuracy and patient management by offering personalized treatment strategies, particularly in cases with high variability (Ge et al. 2024). Similarly, the integration of GraphRAG with LLMs in clinical decision support systems has enhanced decision-making by providing highly contextual and precise predictions, as seen in the study on nephrology (Miao et al. 2024). Other applications include the use of LLMs in psychotherapy to support and automate therapeutic processes (Stade et al. 2024) and in personalized medicine to reduce clinician workload through the automation of administrative tasks (Tripathi et al. 2024). These studies demonstrate the transformational impact of AI in healthcare, setting the stage for further advancements.

The research highlighted in Table II reveal the growing influence of LLMs in healthcare, emphasizing their ability to enhance diagnostic accuracy, automate administrative tasks, and support clinical decision-making across various medical specialties. Our study builds on these advancements by applying RAG-enhanced local LLM specifically to the management of GDM by utilizing a KG framework to capture the complex relationships within GDM-related data. While previous studies, such as the AI model for personalized medicine (Tripathi et al. 2024), and the AI Drug mix and dose Advisor for T2D, highlight the potential of AI to optimize drug dosing and reduce clinician workload by providing personalized treatment recommendations (Nambiar et al. 2024), our approach goes further by directly addressing the need for personalized, adaptive interventions in GDM care. Using local LLMs’ abilities to integrate complex data in real time, our study presents novel contributions in enhancing treatment personalization, making it a significant step forward in AI-driven healthcare applications.

**Table II:** Research Highlighting Representative Use-Cases of AI in Healthcare.

| **Ref.** | **Year** | **Model** | **Key Contributions** |
|---|---|---|---|
| (Ge *et al.* 2024) | 2024 | AI Model for Liver Diseases | Enhanced diagnostic accuracy and patient management tailored for liver diseases. |
| (Ong *et al.* 2024) | 2023 | Clinical Decision Support System | Improved clinical decision-making with RAG-enhanced LLMs, offering precise predictions and treatments. |
| (Miao *et al.* 2024) | 2024 | LLM-RAG for Nephrology | Improved accuracy and reliability in nephrology advice by integrating RAG with LLMs. |
| (Stade *et al.* 2024) | 2024 | LLMs in Psychotherapy | Explored the potential of LLMs to support and potentially automate aspects of psychotherapy. |
| (Tripathi *et al.* 2024) | 2024 | Personalized Medicine AI Model | Demonstrated how LLMs can automate administrative tasks, reducing clinicians' workload from electronic medical records. |

## 3. Prototyping a Proof of Concept

Medical professionals face a challenge in extracting valuable information from the vast amount of data created by scientific activities. Imagine the plight of an over-worked physician in a densely populated part of the Global South. To address this difficulty, computational approaches provide a viable option for generating knowledge representations in the field of medicine. By using these techniques, medical informatics researchers may establish connections between ideas within a particular study and across related inquiries, thus facilitating the development of fresh hypotheses and the progress of research in, for example, the area of GDM intervention suggestions. The method of constructing knowledge representations entails extracting factual information from various portions of scientific literature, which may range from individual phrases to whole paragraphs. For instance, consider the sentence: “Interventions for GDM improve maternal and fetal health outcomes.” Based on this phrase, we can infer several facts: (1) the presence of treatments that specifically focus on GDM; (2) the desired effects of these interventions; and (3) the connection between interventions and the health outcomes of both the mother and the baby.

The research methodology adopted is to develop a Proof-of-Concept (PoC) AI Assistant by creating a comprehensive and parsimonious Knowledge Graph (KG) that depicts the connections between entities related to GDM management by discovering labels or concepts. This relates to the possibility of presenting authoritative medical research findings obtained from peer-reviewed, high-impact publications. The main emphasis at the onset is to identify entities that are important for GDM intervention ideas and highlight the connections between these entities. Figure 2 shows the process flow of our research method, from data collection and entity extraction to the construction and application of the knowledge graph with RAG eventually resulting in a PoC for GDM management. The steps outlined are described in the following sub-sections.

**Figure 2:**
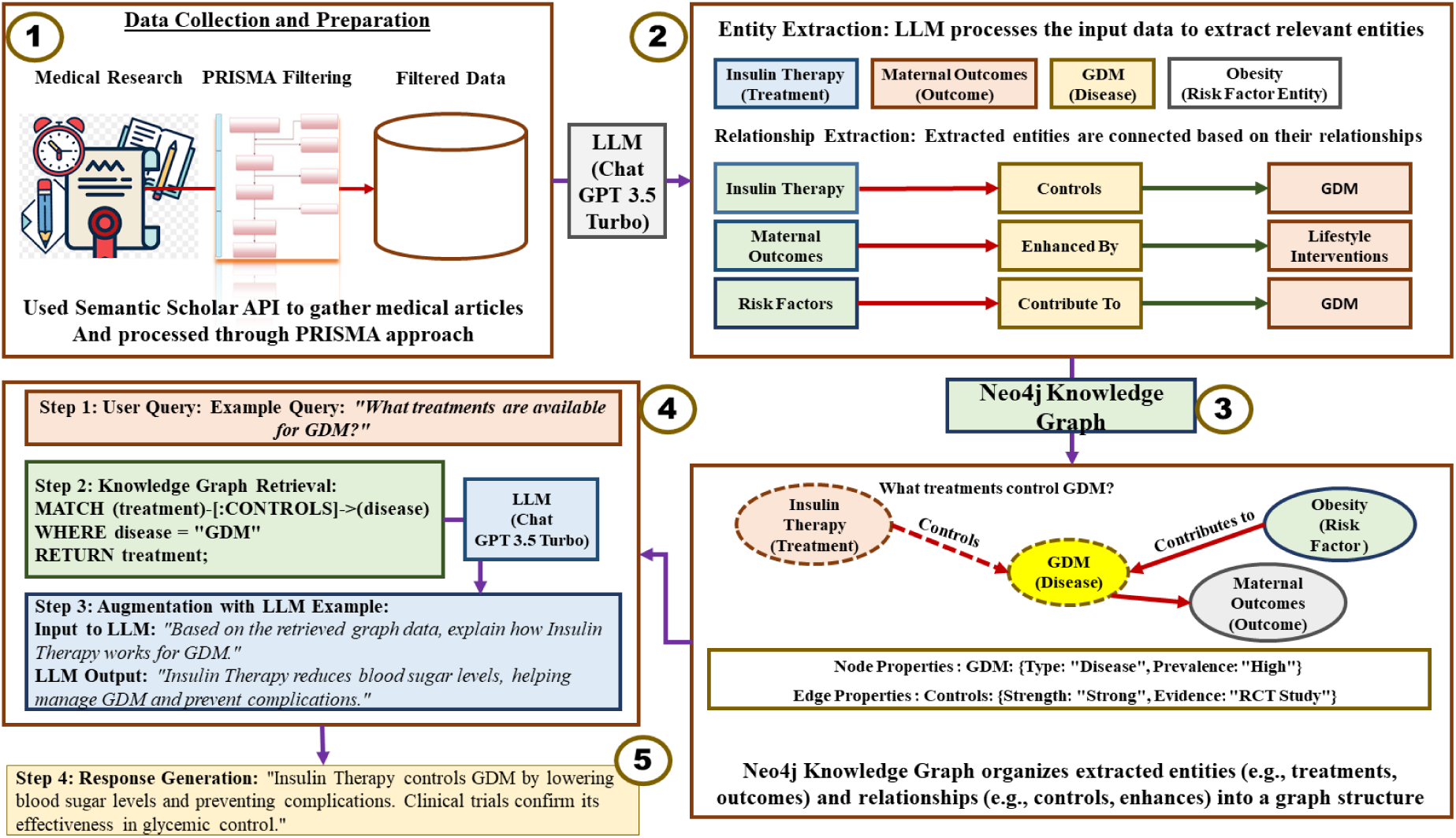
Process flow of the proposed GraphRAG approach, showing data collection, entity extraction, knowledge graph construction, and retrieval-augmented generation for AI-assisted clinical decision support in GDM management.

### 3.1 Data Collection

Data collection for this study was conducted with a systematic search utilizing the Semantic Scholar API, which is well-known for its strong ability to gather academic papers and provide rich metadata coverage (Scholar, 2023). The search criteria used were “gestational diabetes interventions,” to narrow down the query and assure the inclusion of relevant research. The publishing period selected spanned from January 2000 to May 2024, including significant extant as well as current research in GDM interventions. This would enable an in-depth analysis of trends in the diagnosis and treatment of GDM over recent years.

The initial database search produced a total of 2,305 journal articles. These papers underwent a screening process to identify any duplicates and determine their eligibility based on the title and abstract matching “Gestational Diabetes Mellitus”. As a result, the number of records was filtered 1,667. Following that, further screening was conducted to select 1,212 publications for full-text retrieval based on their relevance to “treatments for GDM”. However, a total of 455 publications could not be obtained due to factors such as access limitations or unavailability. Out of the total of 1,212 publications, a detailed evaluation was conducted to determine their eligibility based on certain criteria, including the relevance of the research, the strength of the methodology, and the special emphasis on treatments. All 1,212 papers satisfied these criteria and were included in the review.

Note that while the Semantic Scholar API was chosen for its efficiency and coverage, reliance on a single source could introduce potential biases in the data collection process. The database might not fully represent the global diversity of research on GDM, and the use of specific keywords could lead to a skewed dataset, potentially overlooking alternative perspectives or emerging trends. To address these concerns, future research could consider integrating other databases covering international languages and studies or employing a more comprehensive search strategy to mitigate these biases. For now, our objective was to establish the feasibility of the approach with a PoC.

The filtering approach ensured a strong basis for the next phases of the research. The PRISMA flow diagram shown in Figure 3 depicts the complete account of collecting and filtering data used in this study.

**Figure 3:**
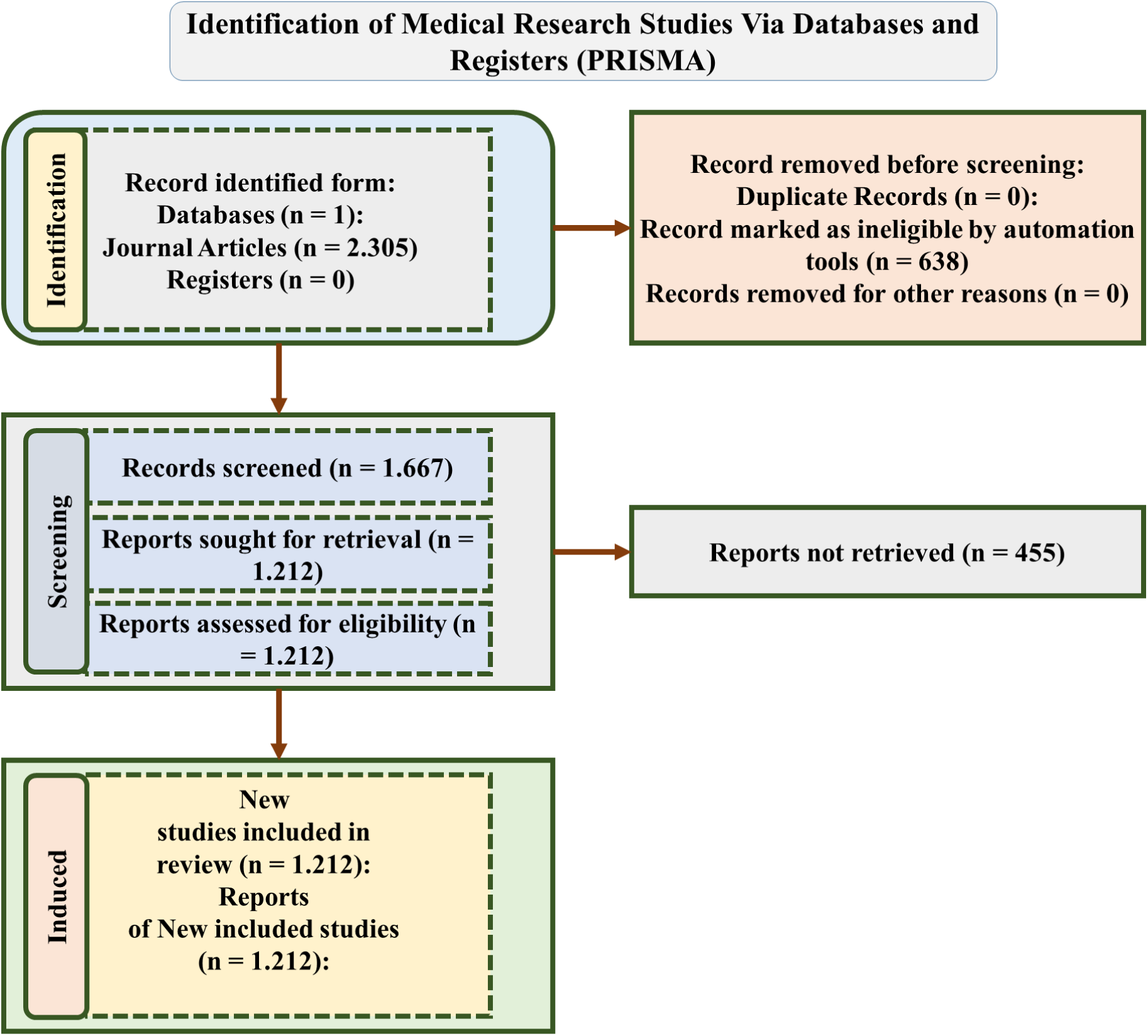
PRISMA flow diagram showing the systematic data collection and filtering process, detailing the stages of identification, screening, eligibility assessment, and inclusion of studies for constructing the knowledge graph.

### 3.2 Entity Extraction

Upon completing the dataset of published GDM research, the next task is to extract precise data points about treatments for GDM from the collection. The entity extraction is performed using the OpenAI API of GPT 3.5 Turbo 16K (OpenAI, 2023), which employs advanced natural language processing (NLP) methods to identify and categorize relevant intervention strategies and their relationships within the literature.

To maximize the precision and relevance of the extraction process, few-shot prompting was utilized. Few-shot prompting is a technique where the API is guided by providing it with a small set of carefully selected examples that illustrate the desired output format. These examples help the model infer patterns, recognize relevant entities, and perform specific tasks with minimal additional supervision. For this task, a handful of representative examples are created focusing on elements of GDM treatments such as the frequency of exercise, nutritional advice, medication doses, and their effects on the health status of both the mother and baby.

The carefully crafted few-shot prompts direct the model’s attention to key aspects of the extracted data while enabling it to adapt to variations in terminology and phrasing across the literature. For instance, equivalent terms like “low carb diet” and “reduced carbohydrate intake” are recognized and consolidated into a single, unified entity ensuring consistency and reducing redundancy. Few-shot prompting is particularly effective in distinguishing between closely related concepts and filtering out irrelevant information. This ensures that the extracted data is both accurate and structured in a way that is conducive to building a knowledge graph (KG).

The application of few-shot prompting not only enhances the precision of the extraction process but also significantly reduces the effort required for manual curation and validation of the data. These actions, categorized as “medical prompt engineering” were carried out by a bioinformatics researcher as part of an experimental PoC. However, for clinical trials, this task would need to be performed or validated by medical specialists to ensure clinical accuracy and compliance. Figure 4 shows the template prompt used for entity extraction.

**Figure 4:**
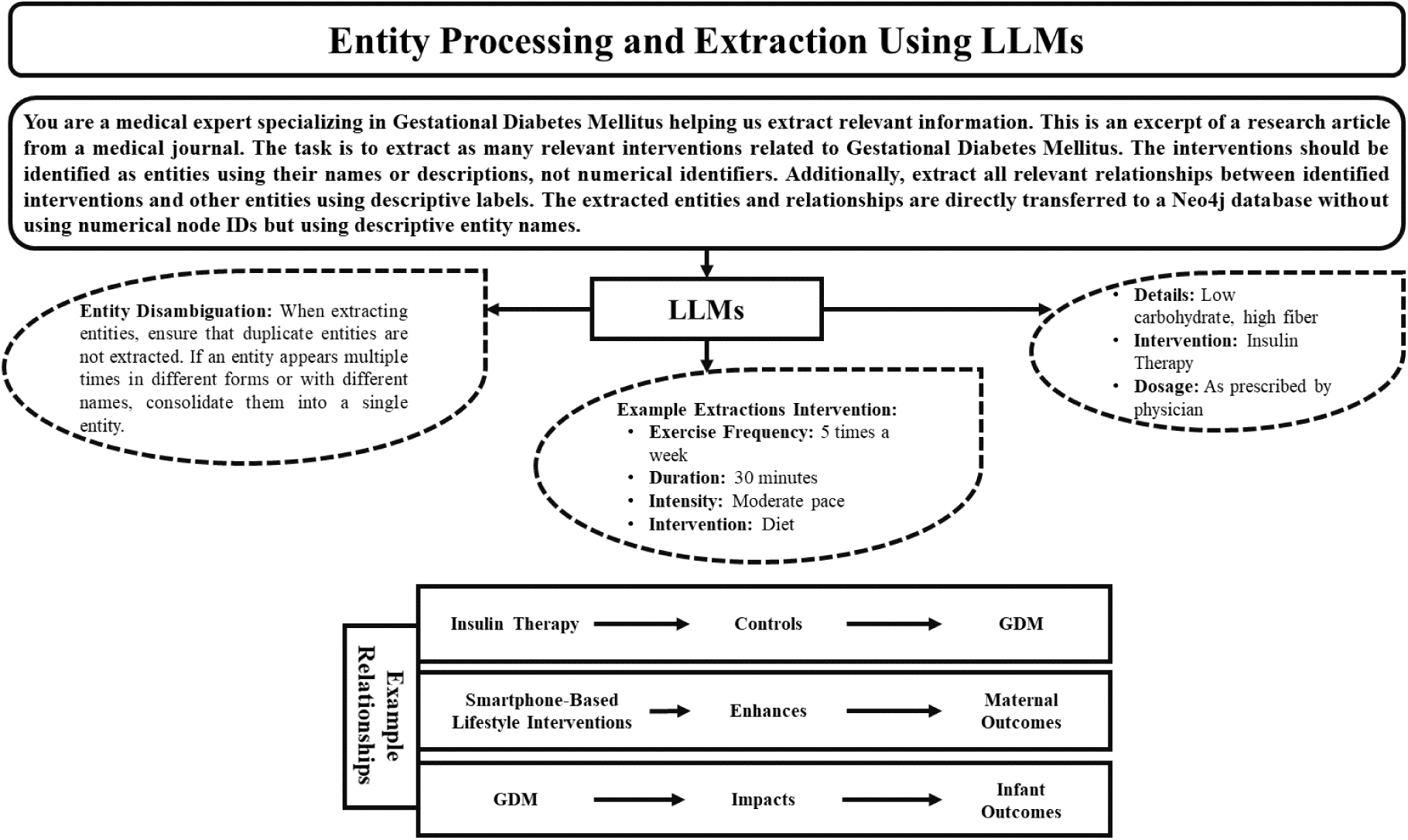
Entity extraction workflow using large language models. The diagram highlights the process of extracting interventions, risk factors, and relationships, ensuring structured and context-aware knowledge representation for GDM management

### 3.3 Construction of the Knowledge Graph

After extracting entities and their relationships from the data, we used Neo4j – an open-source graph database management system – to create a detailed Knowledge Graph (KG) (Trajanoska **et al.** 2023). Neo4j is particularly suited to manage complex and interconnected data structures, making it useful for visualizing the complex relationships in GDM such as interventions, causes, impact, and symptoms (Neo4j, 2023).

We may note that this KG systematically maps out how different treatments, risk factors, and outcomes are interconnected, helping a medical practitioner understand the broader picture of GDM management. The KG is constructed by importing the collected entities and their connections into Neo4j. The system’s advanced query capabilities allow dynamic interaction with the network, facilitating the exploration of complex data connections through clear visualizations. This includes the capacity to track the influence of specific treatments on patient outcomes and understand the relationships among various treatment approaches.

The creation of the KG not only enhances our understanding of GDM interventions but also assists in identifying potentially unexplored regions where further research could be beneficial (Trajanoska et al. 2023). Figure 5 illustrates the interaction between a sample user query, unstructured medical data (including published medical research as well as patient and context-relevant records), and the Neo4j K-Graph using an open-source LLM such as ChatGPT (OpenAI, 2023).

**Figure 5:**
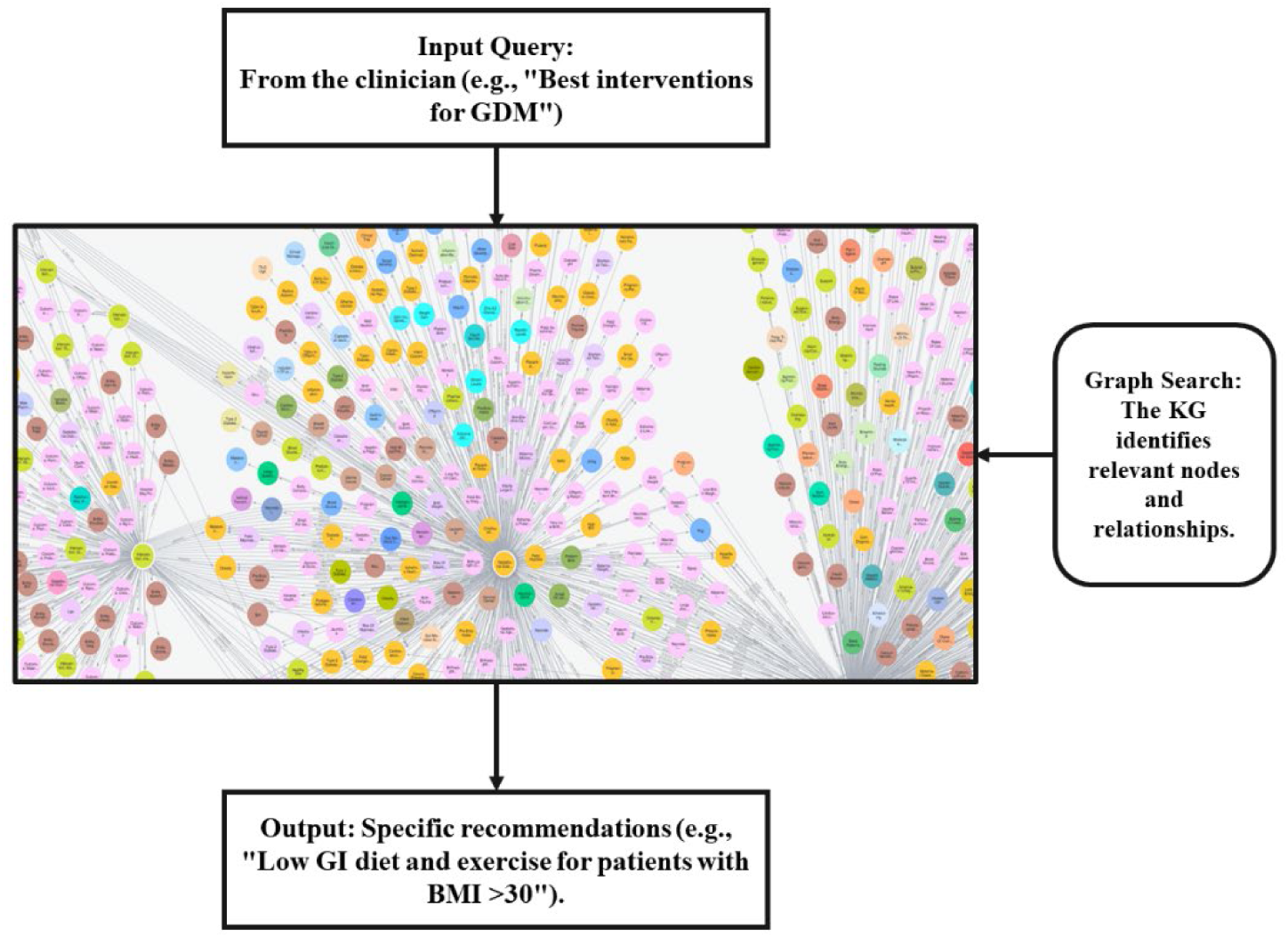
Knowledge Graph-powered clinical support system for GDM. The graph-based search retrieves relevant interventions and relationships, enabling AI-assisted, personalized treatment recommendations.

### 3.4 Knowledge Graph-Based Retrieval-Augmented Generation

To further enhance the utility of the KG, we applied the RAG approach, which combines the strengths of both retrieval-based and generative AI techniques (Hu **et al.** 2024). This approach generates relevant and informative responses based on the data contained within the KG, making it especially useful in clinical decision-making where accuracy and context are paramount.

LLMs, while revolutionary, face significant limitations such as the tendency to produce inaccurate information (hallucinations), the temporal limitations of their knowledge base (including training bias), and difficulties handling complex or domain-specific tasks (Pal **et al.** 2023). The RAG approach helps address these limitations by integrating contextual external data retrieval with LLMs to enhance their performance across various applications, ensuring that responses are not only generated but are grounded in up-to-date, relevant data (Dey **et al.** 2022; Miao **et al.** 2024).

The developed PoC of our Clinical AI Assistant for GDM proceeds in a structured sequence of steps, as visually shown in Figures 6 and 7.

**Figure 6:**
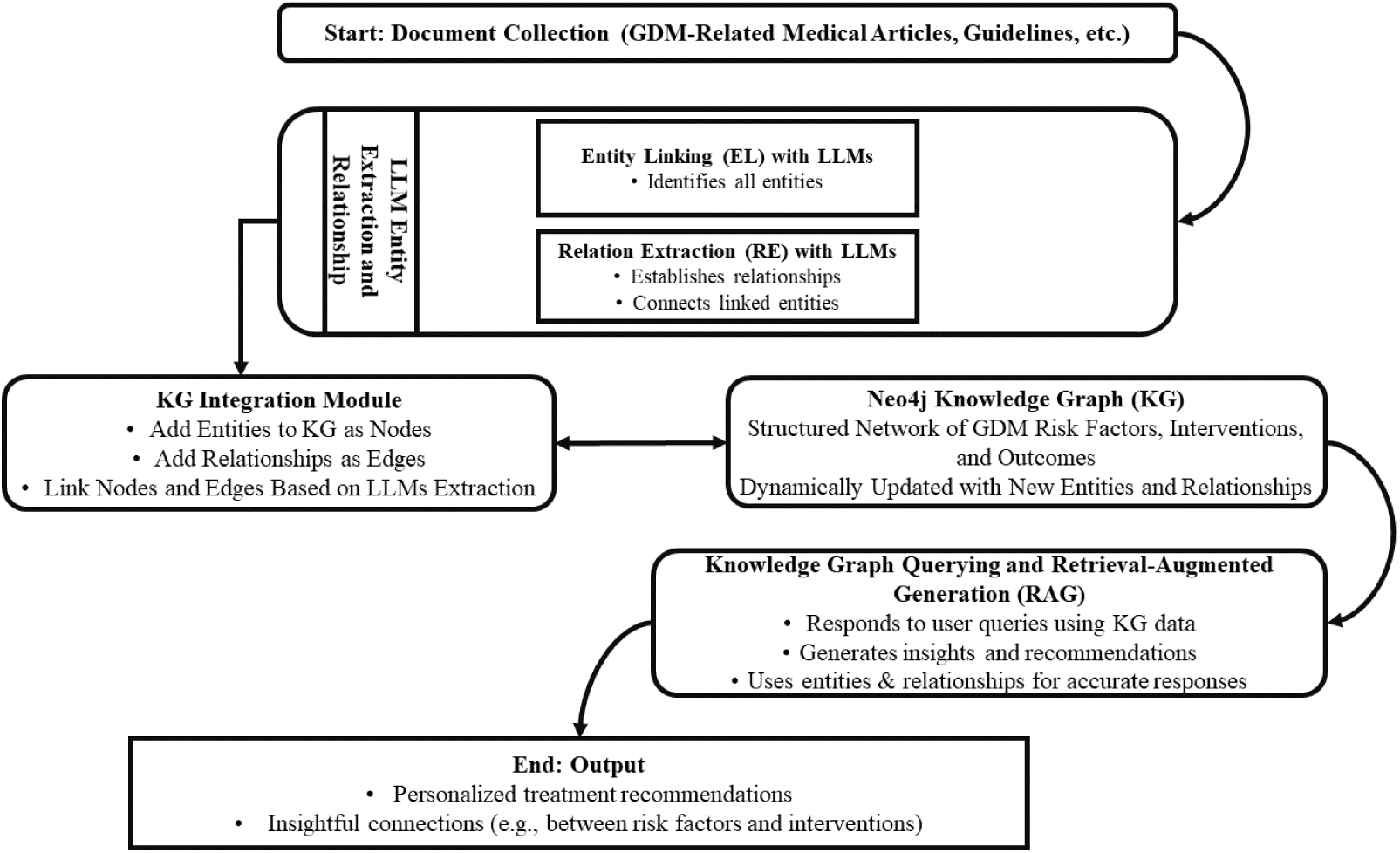
End-to-end process flow of the GraphRAG solution. The pipeline processes medical literature and patient data, integrating them into a structured Knowledge Graph for AI-driven clinical decision support.

**Figure 7:**
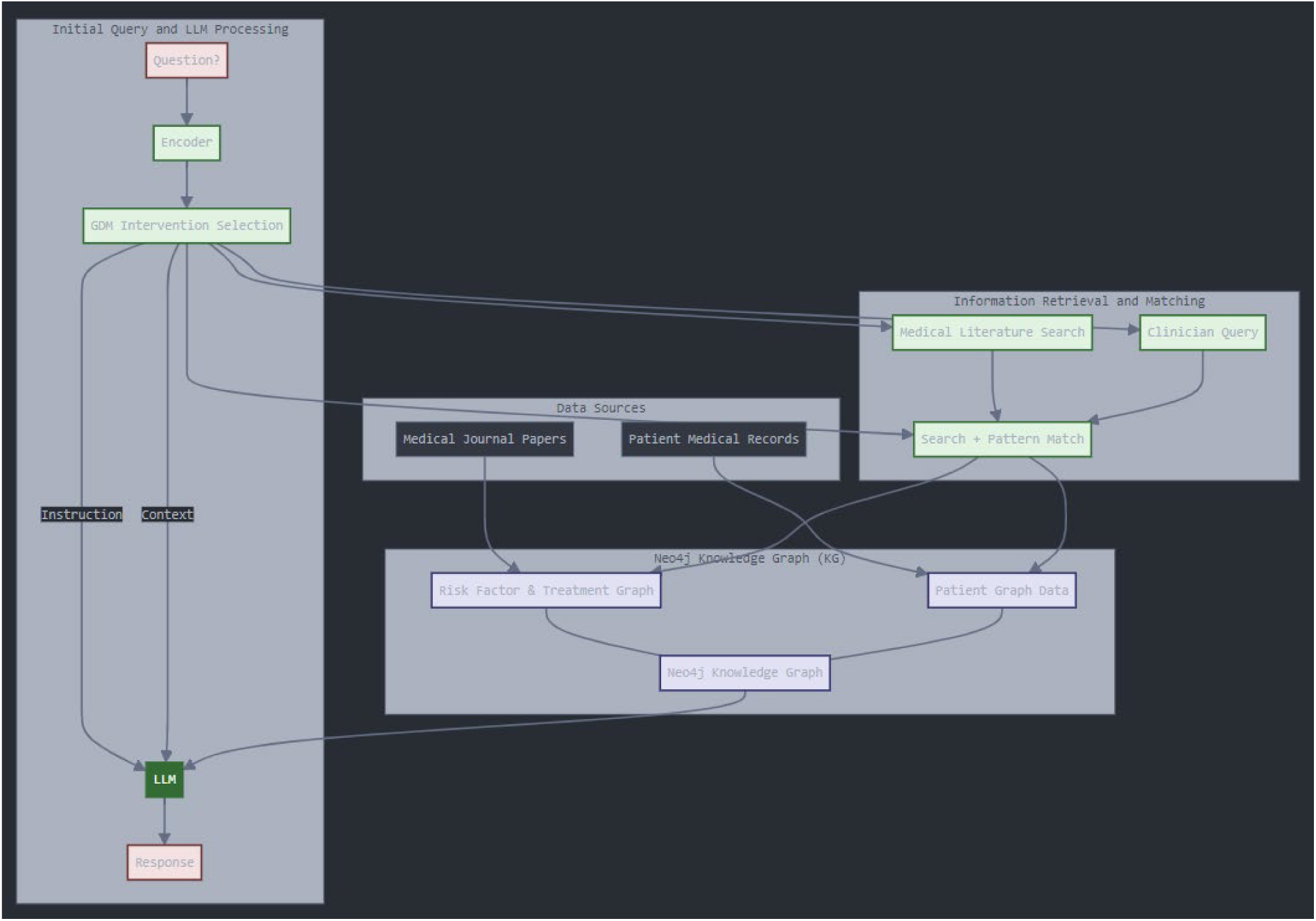
Structured retrieval and response generation process in GraphRAG. The diagram shows how clinician queries interact with medical knowledge sources, pattern matching, and graph-based retrieval to enhance AI-generated responses.

The five-step process increases the likelihood that the responses are both accurate and customized to the specific needs of the user. The system effectively integrates vast amounts of interrelated data, advancing personalized medicine and evidence-based practices in GDM treatment. While Figure 6 and Figure 7 depict a fluid process flow, in reality the art of using RAG and KG entails few-shot training (Stade **et al.** 2024; Tripathi **et al.** 2024) incorporating some trial-and-error in the design of the PoC.

Figure 6 and Figure 7 represent the architecture and processes of our GraphRAG Clinical Decision Support PoC, with clear labels and a logical flow of information. It illustrates the role of AI tools like GraphRAG in healthcare, specifically answering patient questions about GDM or other ailments given the context of domain medical research, patient health records, and other socio-economic considerations. The process flow diagram shows a typical scenario of a numbered step-by-step process from the patient’s question to the final answer. Dynamic interaction is key to advancing healthcare, especially in cases like GDM, where personalized and contextually relevant responses are critical for effective management.

In Figure 7, the top section shows the initial query processing, including the GDM Intervention Selection as per medical guidelines and the LLM components. The middle section illustrates the information retrieval and matching process, incorporating medical literature search and clinician queries. The Neo4j Knowledge Graph section shows how risk factors, treatments, and patient data are integrated. The bottom section labels the two main data sources: Medical Journal Papers and Patient Medical Records with the option of incorporating socio-cultural profiles. The arrow from the Knowledge Graph to the LLM demonstrates how the structured data informs the language model’s responses. Although simplistic,

## 4. Analysis of Results

Beta testing of the PoC in a simulated environment supports the finding that the use of the GraphRAG approach promises to be a powerful tool for the management of GDM. The simulation proceeded as follows. Representative prompts were developed by 5 laypersons (namely, the authors of this paper) and 2 actual General Practitioners (GPs). These prompts were pooled and vetted by 2 specialist doctors before being used to test the efficacy of our proposed GraphRAG local LLM and 3 other widely-used LLMs. Figure 8 and Figure 9 depict a realistic use-case scenario where both healthcare professionals and patients could benefit from the capabilities of the GraphRAG local-LLM.

**Figure 8:**
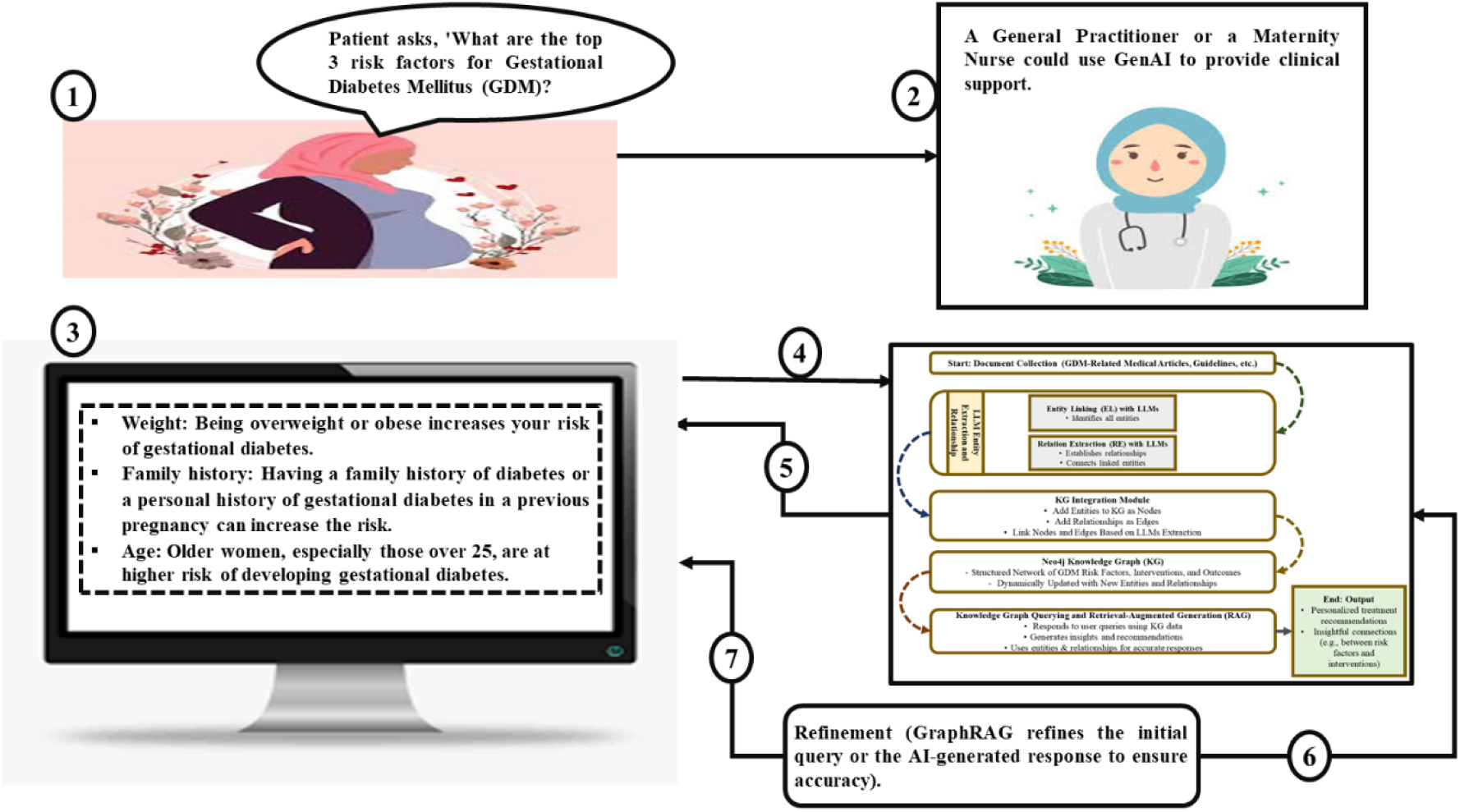
GraphRAG-based Clinical Support System for GDM – Iconographic Representation.

**Figure 9:**
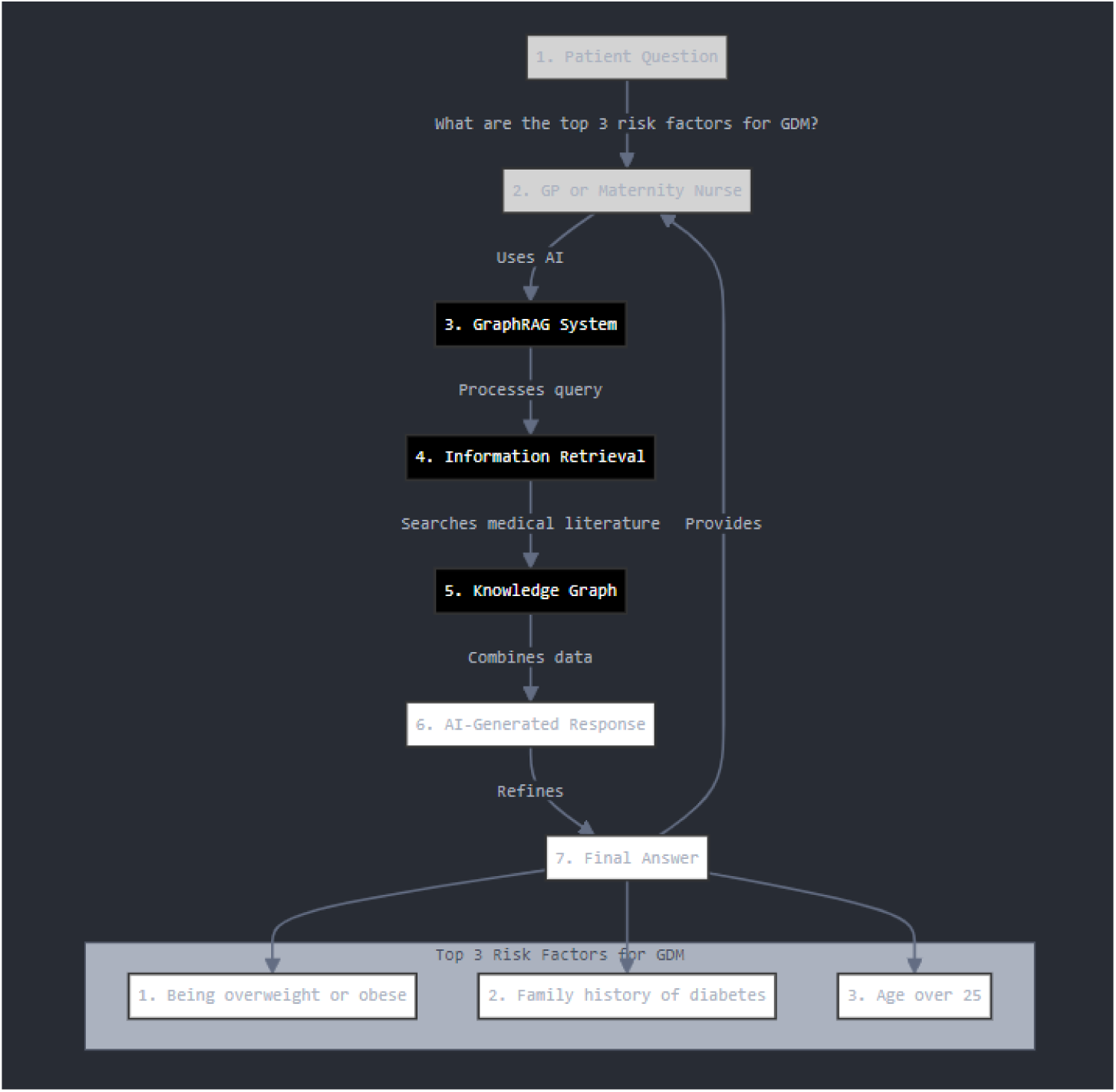
GraphRAG-based Clinical Support System for GDM – Process Flow Diagram.

In this scenario, as illustrated in the iconographic (Figure 8), a patient may ask about the top risk factors for GDM (Step 1). The healthcare professional uses a GenAI clinical support system such as our GraphRAG local-LLM to support GDM patient care (Step 2). The query is processed, and GraphRAG generates a personalized, accurate response (Step 3), listing key GDM risk factors, including weight, family history of diabetes, and age. To continue with the illustration, in steps 4 and 5, the system uses the GDM Intervention selection process and accesses the Neo4j Knowledge Graph, combining information from medical research and patient-specific data. This integration ensures that the AI provides contextually relevant advisories. GraphRAG refines the initial query or the AI-generated response to ensure accuracy and relevance before presenting the final recommendations (Steps 6 and 7).

Figure 9 represents the sequence of processes taken by our GraphRAG Clinical Support System, with clear labels and a logical flow of information. In our illustrated scenario, the medical consultation begins with a patient’s question about risk factors for GDM. A General Physician or Maternity Nurse could use the GraphRAG system to probe for an expert medical opinion. The system processes the query through information retrieval and a knowledge graph. An AI-generated response is generated by what we architect as a local LLM since contextual information is the basis of its response. The healthcare professional provides the advise to the patient as a value-added advisory. Here, the top 3 risk factors for GDM are listed at the bottom of the diagram.

This refinement process is critical for customizing the response based on the patient’s profile and the clinician’s query, ensuring that the advice is not only accurate but also highly personalized. In this example, the GP receives an AI-generated summary of the most critical GDM risk factors based on the query. The GraphRAG system can generate a response that is both precise and aligned with up-to-date medical knowledge. This capability reduces the likelihood of hallucination or bias, providing the patient with reliable, evidence-based advice while supporting the GP in her decision-making process.

The versatility of the GraphRAG model extends beyond patient interactions. Healthcare professionals can also use it to address specific clinical questions, such as determining the most effective screening tests for GDM or assessing the effectiveness of particular interventions. This provides support to both healthcare practitioners and patients, highlighting the significant potential of GraphRAG in clinical settings, and enhancing evidence-based practices in the management of GDM.

Our analysis of the efficacy of the PoC is detailed in the following sub-sections. In 4.1, Response to Layperson Prompts assesses how effectively the system simplifies complex information for the general public. We next analyze responses to prompts from two General Practitioners (GPs) to evaluate the model’s clinical relevance in 4.2. And in 4.3, the Comparative Analysis of Model Performance uses standard evaluation metrics to assess GraphRAG’s performance against other commonly adopted LLMs such as Chat GPT, Claude, and Mistral.

### 4.1 GraphRAG local-LLM Response to Layperson Prompts

The GraphRAG system provides valuable insights into several aspects of GDM management that are easy for the general public to understand. For example, one typical question might be: “What are the top 3 factors that determine the risk of gestational diabetes?” Detailed system responses to this and other prompts are presented in Table III.

**Table III:** Responses to Layperson Prompts.

| **Prompt** | **Response** |
|---|---|
| *What are the top 3 factors determining gestational diabetes risk?* | **Weight**: Being overweight or obese increases your risk of gestational diabetes.<br>**Family history**: Having a family history of diabetes or a personal history of gestational diabetes in a previous pregnancy can increase the risk.<br>**Age**: Older women, especially those over 25, are at higher risk of developing gestational diabetes. |
| *How can we safeguard a baby’s health and wellness for gestational diabetes patients?* | Monitor blood sugar levels, follow a healthy meal plan, engage in regular physical activity, take prescribed medications, attend prenatal check-ups, and follow healthcare guidance. |
| *What nutrition is recommended to prevent gestational diabetes?* | Eat a balanced diet high in fiber and low in added sugars and unhealthy fats. Include fruits, vegetables, whole grains, lean proteins, and healthy fats. |
| *Are gestational diabetes patients more likely to have a cesarean or normal delivery?* | Women with gestational diabetes are more likely to have a cesarean delivery due to risks like macrosomia, shoulder dystocia, and birth trauma. |
| *What neonatal screening should a couple with a family history of diabetes undergo before planning to have a baby? Are there precautions they could take?* | Consider genetic counseling and testing. Preconception counseling can help identify risks and precautions, with personalized plans developed alongside healthcare professionals |

To summarize the key takeaways from the above layperson-origin queries and the corresponding model-generated responses:

#### Summary of Responses on Gestational Diabetes to Layperson-Oriented Prompts

- **Risk Factors and Prevention:** The main factors influencing the risk of gestational diabetes include being overweight or obese, having a family history of diabetes, or being over the age of 35. Monitoring and controlling weight, understanding genetic traits, and being aware of age-related risks are essential to avoiding GDM.
- **Health and Wellness of the Child:** Ensuring the child’s health and wellness involves carefully monitoring blood glucose levels, following a nutritious diet, participating in regular physical exercise, taking prescribed medications as necessary, and attending routine prenatal examinations. This approach helps maintain optimal health for both mother and child.
- **Nutrition Recommendations:** A well-balanced diet rich in fiber and low in added sugars and harmful fats is recommended to reduce the risk of gestational diabetes. Consuming a variety of fruits, vegetables, whole grains, lean proteins, and healthy fats is crucial. Proper nutrition supports better glucose metabolism and overall health.
- **Cesarean Delivery:** Women with gestational diabetes are more likely to have a cesarean delivery due to the increased risk of complications such as macrosomia (large birth weight), shoulder dystocia, and birth trauma. Understanding these risks enables better planning and management of delivery outcomes.

### 4.2 GraphRAG local-LLM Response to the General Practitioner (GP) prompts

The use of realistic prompts from medical practitioners such as physicians and maternity nurses provides a robust framework for exploring the capabilities of the GraphRAG system for addressing specific clinical questions. This approach not only supports the accuracy and relevance of the AI-generated responses but also mimics the practical querying behavior seen in clinical settings. Section 4.2 discuss one of the prompts as an example to familiarize readers with the content, followed by a detailed presentation of prompts from General Practitioner 1 (GP 1). For example, consider GDM’s diagnostic process. The prompt given to the GraphRAG system was: “How do I diagnose gestational diabetes?” The response from the GraphRAG system suggests the use of a glucose challenge test followed by a glucose tolerance test if needed, detailing the procedures to be taken. This example revealed the system’s ability to provide precise and actionable medical advice.

#### 4.2.1 Prompts from GP 1 and GraphRAG local-LLM Responses

This subsection discusses the responses provided by the GraphRAG system to the prompts suggested by GP 1, which are relevant to understanding and managing GDM effectively. These responses are detailed in Table IV and are summarized below:

**Table IV:**
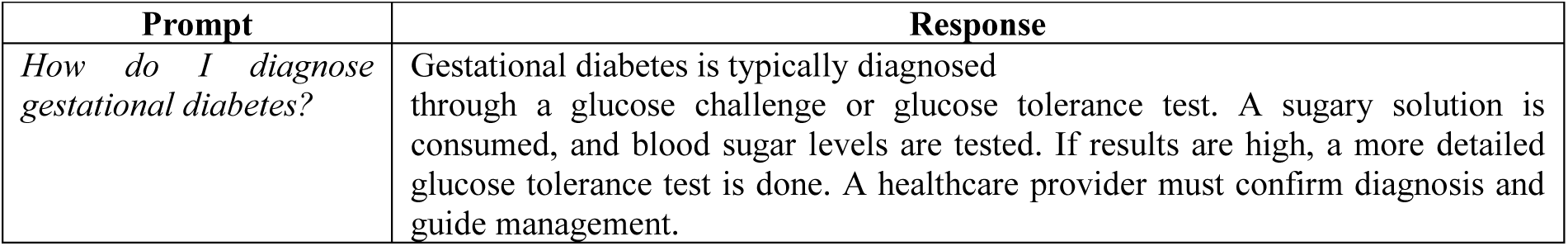
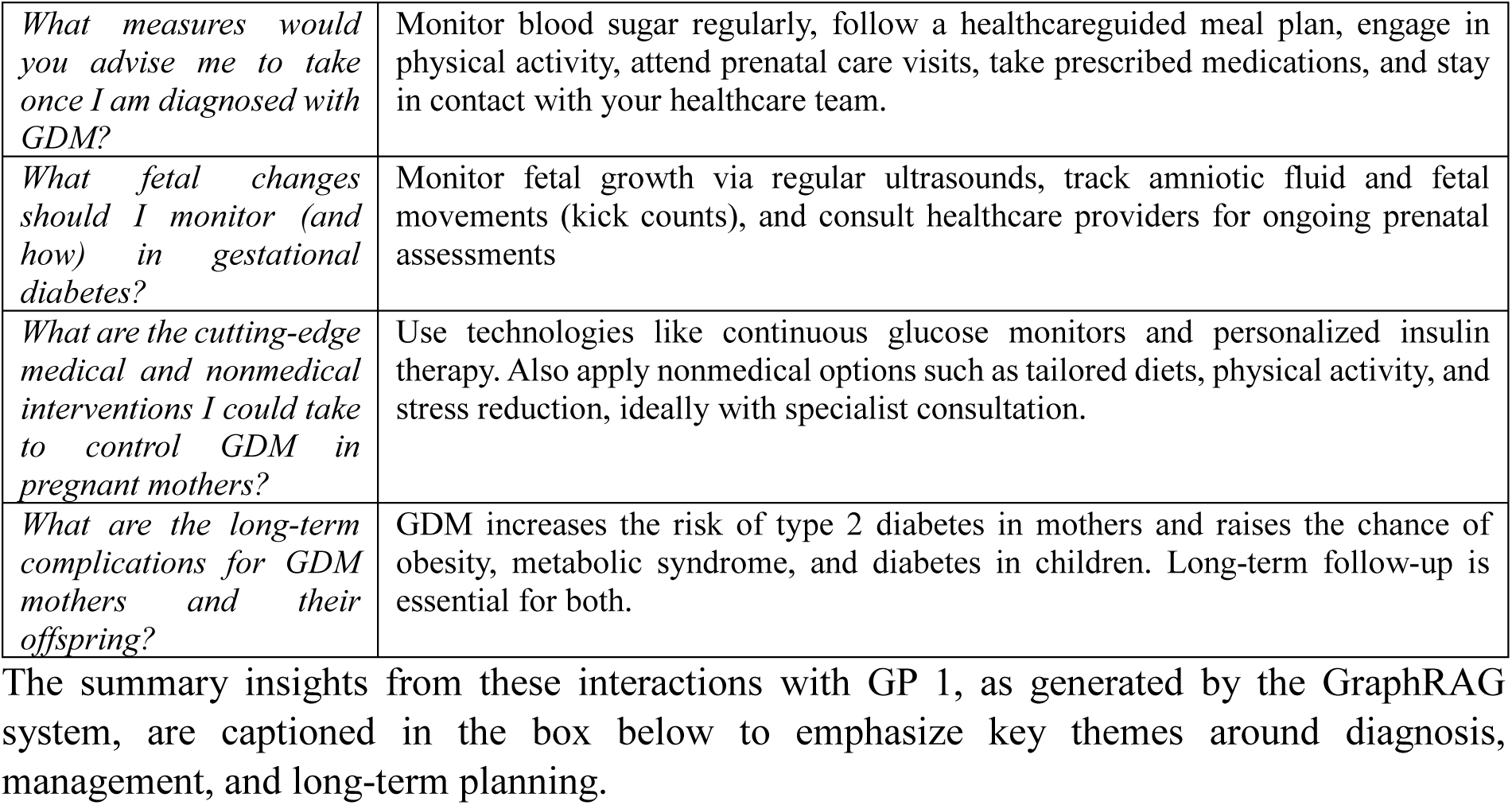
Responses to Prompts from GP 1.

| Prompt | Response |
|---|---|
| *How do I diagnose gestational diabetes?* | Gestational diabetes is typically diagnosed<br>through a glucose challenge or glucose tolerance test. A sugary solution is consumed, and blood sugar levels are tested. If results are high, a more detailed glucose tolerance test is done. A healthcare provider must confirm diagnosis and guide management. |
| *What measures would you advise me to take once I am diagnosed with GDM?* | Monitor blood sugar regularly, follow a healthcareguided meal plan, engage in physical activity, attend prenatal care visits, take prescribed medications, and stay in contact with your healthcare team. |
| *What fetal changes should I monitor (and how) in gestational diabetes?* | Monitor fetal growth via regular ultrasounds, track amniotic fluid and fetal movements (kick counts), and consult healthcare providers for ongoing prenatal assessments |
| *What are the cutting-edge medical and nonmedical interventions I could take to control GDM in pregnant mothers?* | Use technologies like continuous glucose monitors and personalized insulin therapy. Also apply nonmedical options such as tailored diets, physical activity, and stress reduction, ideally with specialist consultation. |
| *What are the long-term complications for GDM mothers and their offspring?* | GDM increases the risk of type 2 diabetes in mothers and raises the chance of obesity, metabolic syndrome, and diabetes in children. Long-term follow-up is essential for both. |

##### Key Insights from System Responses to GP1 Prompts

- **Diagnosis of Gestational Diabetes:** The system explains the use of glucose challenge and tolerance tests to diagnose GDM, emphasizing the importance of initial and follow-up testing procedures.
- **Management Measures Post-Diagnosis:** It suggests regular monitoring of blood sugar levels, dietary management, physical activity, and medication adherence. The importance of prenatal care and communication with the healthcare team are also highlighted.
- **Monitoring Fetal Changes:** Recommendations include regular ultrasounds to assess fetal growth and the monitoring of the baby’s movements, reflecting comprehensive maternal and fetal healthcare.
- **Cutting-edge Interventions:** Discusses the integration of modern technologies like continuous glucose monitoring and personalized insulin therapy alongside non-medical interventions such as nutrition and exercise plans.
- **Long-term Complications:** Outlines the increased risks of type 2 diabetes, obesity, and metabolic syndrome for both mothers and their offspring, stressing the need for long-term follow-up care.

#### 4.2.2 Prompts from GP 2 and GraphRAG Local-LLM Responses

Upon assessing the responses generated by the GraphRAG system to the prompts suggested by GP 1, we further explored the robustness of the system by introducing more clinically focused prompts from GP 2. This section discusses the interactions between GP 2 prompts and the GraphRAG responses, covering a wider scope of GDM management. The responses, shown in Table V, are useful for enhancing our understanding of how GenAI solutions can effectively address complex clinical scenarios in GDM management.

**Table V:** Responses to Prompts from GP 2.

| Prompt | Response |
|---|---|
| *What basic screening tests for GDM should I perform and monitor regularly?* | Common tests for screening and monitoring GDM include:<br>1. **Glucose Challenge Test (GCT)** – a one-hour blood test after consuming a sugary drink.<br>2. **Oral Glucose Tolerance Test (OGTT)** – a follow-up fasting test with multiple blood draws.<br>3. **Glycated Hemoglobin (A1C)** – reflects average glucose levels over past months.<br>4. **Regular Blood Glucose Monitoring** – daily checks using a glucometer.<br>Consult a healthcare provider for tailored testing schedules |
| *On the balance, does medical evidence suggest a greater genetic influence of the father or mother for DM?* | Both parents can genetically influence the risk of DM. The extent varies by DM type and individual genetic profiles. Environmental factors also play a key role. |
| *If the patient and her husband decide to go ahead with starting a family, what medical interventions can I prescribe to alleviate the risk of GDM?* | Medical interventions include:<br>1. Healthy diet and lifestyle habits.<br>2. Regular blood glucose monitoring.<br>3. Medications such as metformin if required.<br>4. Referral to a nutritionist or diabetes educator.<br>5. Recommendations should be personalized by the provider. |
| *Can proper oral and dental care alleviate GDM during pregnancy? What are some effective measures?* | There is no direct link between oral care and GDM<br>prevention. However, good dental hygiene supports overall pregnancy health. Recommendations include brushing twice daily, flossing, and routine dental visits. |
| *Are there promising stem-cell or epigenetic treatments on the horizon that could ease my patient’s hesitance to start a family?* | Stem-cell and epigenetic research shows potential but is still in the early stages. Patients should consult reproductive specialists for the latest updates and personalized advice |

The most relevant insights extracted from these responses—ranging from diagnostics to novel therapies—are summarized in the Box below to highlight key clinical themes.

##### Key Insights on System Responses to GP2 Prompts

- **Basic Screening and Monitoring Tests:** GraphRAG recommends the Glucose Challenge Test (GCT) at 24–28 weeks for initial screening, followed by the Oral Glucose Tolerance Test (OGTT) if needed. The Glycated Hemoglobin (A1C) test offers long-term glucose averages. Regular daily blood glucose monitoring is advised using a glucometer.
- **Genetic Influence on Diabetes Mellitus:** Both parents contribute to genetic risk for DM, influenced by individual genetic makeup and environmental factors. The impact varies by DM type.
- **Medical Interventions to Reduce GDM Risk:** Suggested interventions include dietary and lifestyle changes, regular glucose monitoring, medications like metformin when necessary, and referrals to nutritionists or diabetes educators.
- **Oral and Dental Care during GDM:** GraphRAG found no direct medical link between oral care and GDM management but supports good oral hygiene for overall prenatal health—highlighting its reduced tendency to hallucinate compared to standard LLMs.
- **Stem-Cell and Epigenetic Therapies:** GraphRAG recognizes the potential of these treatments for reproductive health, including GDM, but stresses they are still experimental and not yet clinically available.

The responses to both the layman and the GP personas clinically demonstrate the versatility and depth of the GraphRAG model. When addressing layman prompts, the model effectively simplifies complex medical information, making it accessible and understandable for the general public. It provides clear and practical guidance on key aspects of GDM, such as risk factors, health and wellness, and dietary recommendations. For GP 1, the model delivers precise and actionable advice on diagnosing and managing GDM, including specific tests and interventions. These responses highlight the abilities of the model to support healthcare professionals in making informed clinical decisions.

Beyond initial evaluations, we introduced more clinically focused prompts from GP 2 to further ascertain the robustness of the model. The interactions with GP 2 prompts reveal the capability of the GraphRAG model to handle a wide range of medical queries, including advanced topics like genetic influences on DM and emerging treatments such as stem-cell and epigenetic therapies. These responses highlight the effectiveness of the model in providing healthcare professionals with accurate, contextually relevant information that enhances decision-making processes in clinical settings.

### 4.3 Evaluation Metrics for GraphRAG and LLMs in GDM Use Case

This aspect of the PoC beta test used standard evaluation methods to measure how well the GraphRAG-enabled local LLM benchmarks with 3 widely-used LLMs when presented with prompts on managing GDM. The metrics used assess how well each model provides clinically relevant, precise, and interpretable information for healthcare providers and patients. The selected metrics were derived from similar studies (Ngo **et al.** 2024; Xiong **et al.** 2024). Namely, the Relevance Score, BLEU Score, Jaccard Similarity, BERT Score, and METEOR, each serving a distinct purpose in assessing model performance.

Evaluation Metrics for AI models used in healthcare require much stricter testing than general-purpose language models. For general language models, it’s often enough if they are fluent and make sense. The selected metrics ensure that AI-generated responses maintain clinical accuracy, contextual relevance, technical precision, and interpretability. Since medical decision-making relies on factual correctness and contextual understanding, these metrics help measure how well AI-generated outputs align with validated medical knowledge and structured clinical guidance.

#### 4.3.1 Comments on the Evaluation Metrics

- **Relevance Score**: This metric quantifies how closely the model’s responses align with the specific informational needs of healthcare providers and patients. Given the high-stakes nature of medical decision-making, an AI system must provide responses that are not only factually correct but also directly applicable to the user’s query. A higher relevance score indicates better adaptability of the model in addressing medical concerns.
- **BLEU Score:** The Bilingual Evaluation Understudy (BLEU) Score measures how well the model replicates validated medical text structures. In clinical AI, this ensures that responses maintain the clarity, precision, and terminology of authoritative sources, reducing the risk of ambiguous or misleading answers. This is particularly critical in healthcare settings where textual precision directly influences decision-making.
- **Jaccard Similarity**: This metric evaluates the overlap of key medical concepts and terminologies between model-generated responses and reference texts. A high Jaccard Similarity score signifies that the AI system preserves essential medical terminology, which is crucial in GDM management. Ensuring terminology consistency helps in preventing misinterpretations that could affect clinical decision-making.
- **BERT Score:** Unlike BLEU and Jaccard Similarity, the BERT Score assesses contextual accuracy by measuring semantic similarity between AI-generated responses and expert-validated texts. This metric is particularly useful in evaluating responses that require complex medical explanations, where contextual relationships between terms such as **insulin resistance** and “glucose intolerance” need to be correctly understood.
- **METEOR**: The Metric for Evaluation of Translation with Explicit ORdering (METEOR) assesses fluency and coherence in AI-generated responses. In healthcare applications, responses must not only be factually correct but also well-structured and easy to interpret. This metric captures how well the AI system formulates responses that facilitate comprehension for both medical professionals and patients.

#### 4.3.2 Performance Benchmarking

To ensure a fair and complete comparison, the models are tested using a standardized set of clinical queries that cover various aspects of GDM-related decision-making. These queries included layperson prompts to assess how well the model simplifies medical information, specialized clinician queries to evaluate its ability to respond to expert-level medical questions, and evidence-based medical prompts to measure its alignment with established clinical guidelines. Each model’s performance is assessed using the metrics above, with results analyzed for statistical significance to validate observed differences.

The following subsection details the benchmarking results, highlighting how GraphRAG performs compared to widely used LLMs such as ChatGPT, Claude, and BioMistral.

To assess the effectiveness of language models in managing GDM, we have conducted a comparative evaluation of GraphRAG, BioMistral, ChatGPT, and Claude. Each of these models has distinct characteristics and strengths in medical AI applications. The goal of this evaluation is to determine their ability to generate accurate, contextually relevant, and clinically actionable responses. The analysis is conducted using established evaluation metrics, including Relevance Score, BLEU Score, Jaccard Similarity, BERTScore, and METEOR.

#### 4.3.3 Overview of Language Models Evaluated

BioMistral is a specialized model designed for medical applications, optimized for clinical text processing with a reduced number of parameters. Its design allows for high efficiency in patient-centered interactions, making it particularly effective in direct healthcare applications. ChatGPT, on the other hand, is a general-purpose LLM capable of handling diverse queries, but its broad focus sometimes results in responses that lack the specificity required for complex medical cases such as GDM management. Claude shares many similarities with ChatGPT but operates on a different architecture, offering coherent and contextually rich responses while still facing limitations in highly specialized medical contexts. GraphRAG is specifically designed for medical environments, integrating retrieval-augmented generation techniques to improve the accuracy and contextual depth of responses. By leveraging structured knowledge retrieval and generative AI, GraphRAG intends to provide healthcare professionals with precise, actionable insights. A summary of the general strengths and limitations of each model is provided in Table VI.

**Table VI:** Comparison of Benchmark Language Models.

| **Model** | **Strengths** | **Limitations** |
|---|---|---|
| BioMistral (Labrak *et al.* 2024) | Optimized for medical text processing, efficient for direct patient interactions | Limited general knowledge outside medical contexts |
| ChatGPT (OpenAI, 2023) | Versatile, capable of handling diverse medical queries | General-purpose model, lacks domain-specific precision |
| Claude (Anthropic, 2023) | Coherent and contextually rich responses, similar to ChatGPT in general use cases | Not specialized for highly technical medical discussions |
| GraphRAG | Retrieval-augmented approach enhances medical accuracy, customized for clinical applications | Proof-of-concept stage, requires further clinical validation |

#### 4.3.4 Performance Evaluation Using Standard Metrics

The evaluation of the benchmark models is conducted using multiple metrics to assess accuracy, terminology preservation, contextual relevance, and coherence. The bar chart in Figure 10 presents a comparative analysis of the average performance across these metrics, offering insights into how well each model performs in GDM-related healthcare prompts.

**Figure 10:**
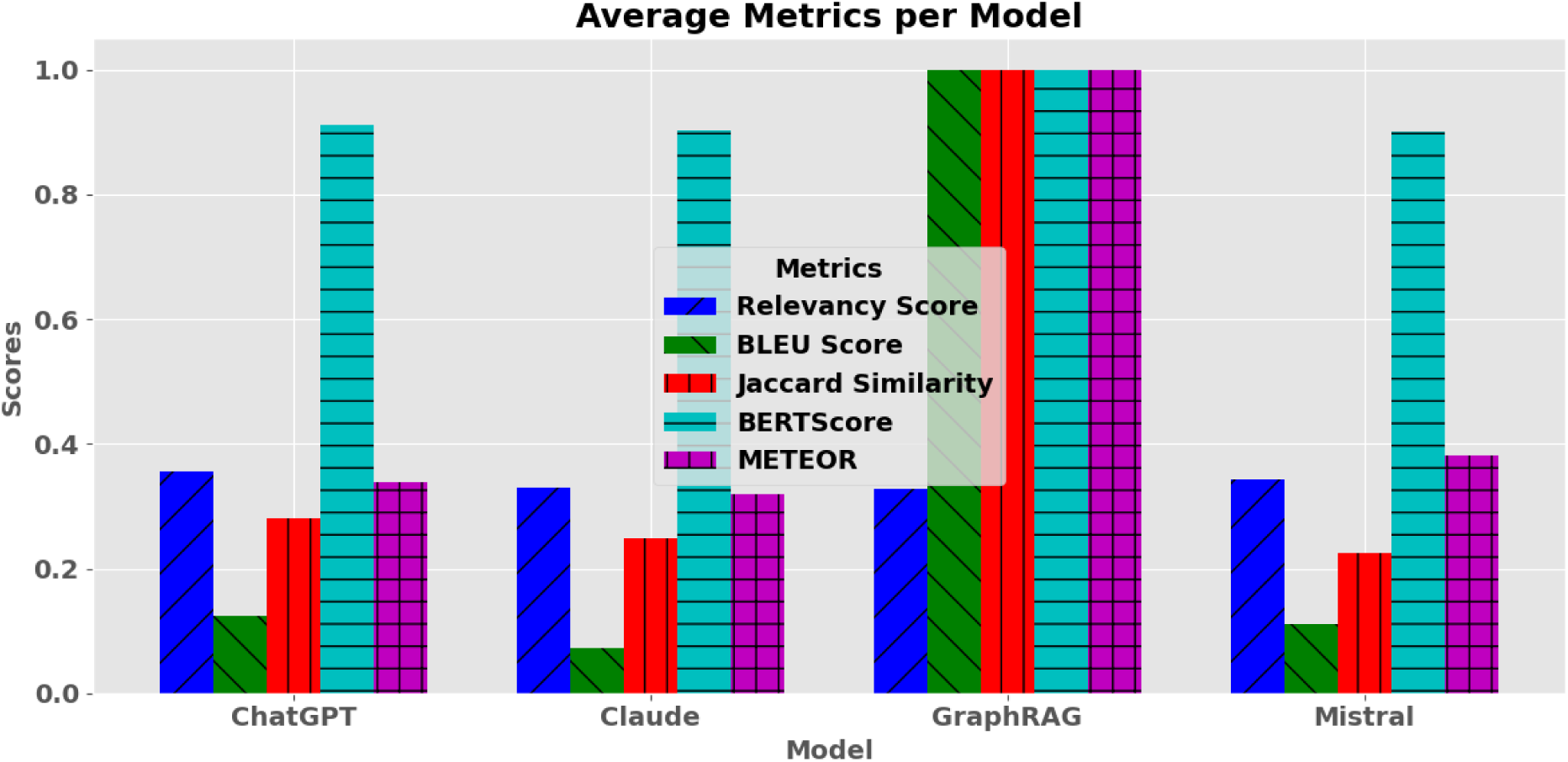
Visual benchmarking of model performance across evaluation metrics.

The radar charts in Figure 11 further provides a multidimensional perspective by plotting each metric along separate axes, enabling a more comprehensive assessment.

**Figure 11:**
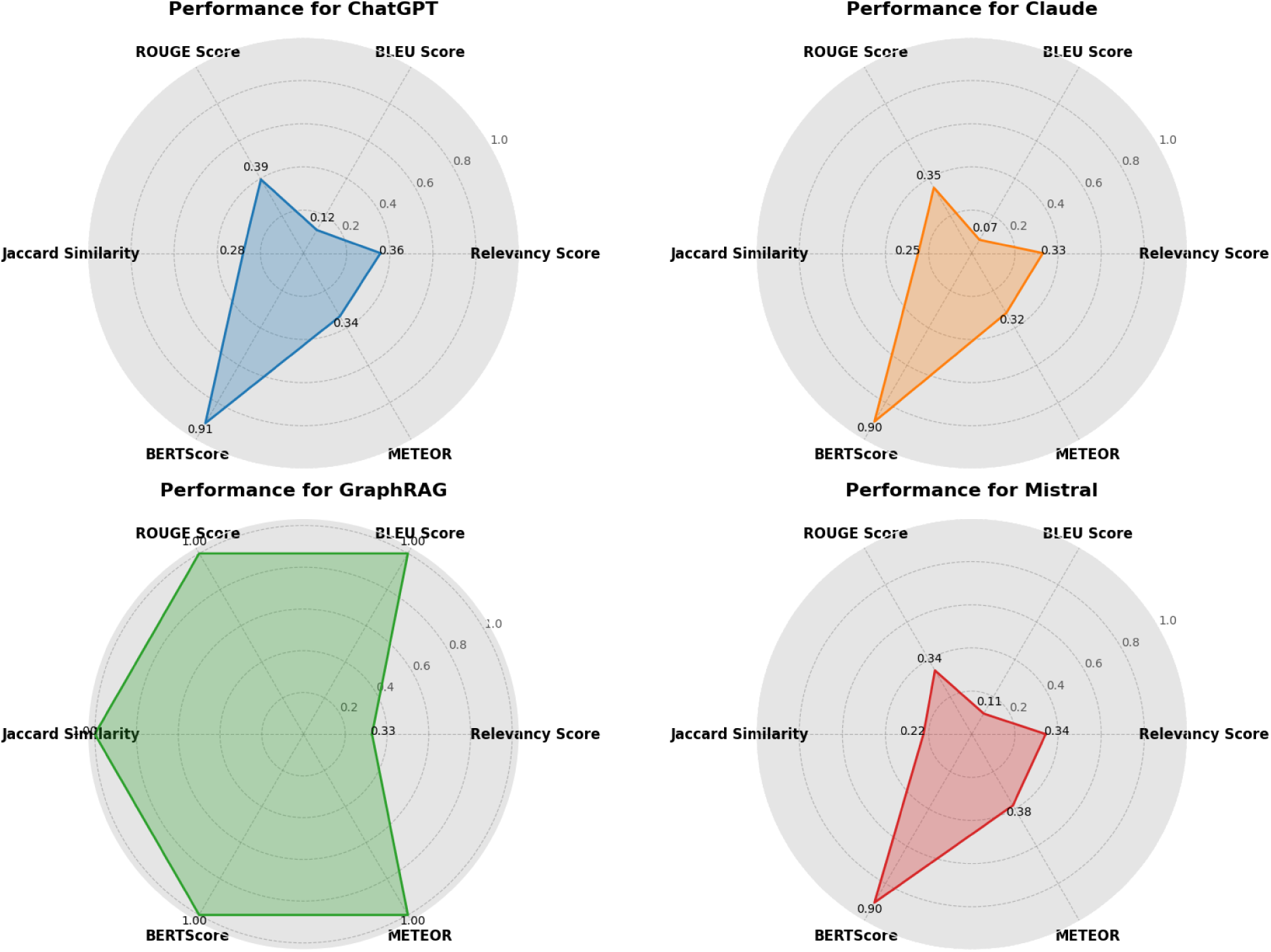
Radar chart visualizing model performance across multiple dimensions.

##### Key Observations from Model Performance

GraphRAG performs comparably to other models in Relevance Score, ensuring that responses align with the needs of healthcare providers managing GDM. Its high BLEU Score shows strong precision in replicating validated medical knowledge, ensuring the clarity and accuracy of clinical communications. The model consistently outperforms others in Jaccard Similarity, maintaining a high level of consistency in medical terminology, which is crucial for technical discussions related to GDM. A high BERTScore reflects its deep contextual understanding, particularly beneficial for generating responses customized to complex patient care situations. The METEOR score highlights its ability to produce well-structured, coherent responses that offer actionable insights to healthcare professionals.

##### Implications for AI-driven Healthcare Solutions

The evaluation results suggest that GraphRAG holds strong potential for use in clinical decision support systems, in this use-case – GDM management. By balancing retrieval-based knowledge integration with generative AI, it effectively combines the strengths of knowledge-based and DL models. While BioMistral remains an efficient choice for patient-centered interactions, it lacks the broader adaptability needed for complex decision-making scenarios. ChatGPT and Claude, while offering versatility, struggle with domainspecific precision, reinforcing the need for medical-specialized models such as GraphRAG. This assessment highlights the feasibility of GraphRAG in advancing AI-driven healthcare solutions. The model’s balanced performance across key metrics suggests its potential for improving clinical decision support, enabling more precise, contextually relevant, and interpretable AI-generated recommendations in healthcare settings.

### 4.4 Effectiveness of the GraphRAG local-LLM for GDM Clinical Support

Advancements in AI-driven healthcare have led to the development of more specialized and context-aware models, with GraphRAG showing significant improvements over general-purpose LLMs in clinical decision support. Unlike conventional models, GraphRAG integrates RAG and KG structuring, allowing it to provide more precise, evidence-based, and contextually relevant insights customized for GDM management. By utilizing structured medical knowledge and continuously updated literature, GraphRAG enhances interpretability and reliability in clinical applications.

- **Clinical Utility and Contextual Relevance**: The ability of GraphRAG to deliver highly relevant responses in GDM management stems from its structured retrieval mechanism and domainspecific training. Traditional LLMs such as ChatGPT and Claude, while effective in general contexts, lack the depth required for specialized medical applications. In contrast, GraphRAG aligns its responses with validated clinical guidelines, ensuring that recommendations are not only accurate but also applicable in real-world scenarios. The model’s enhanced performance in structured evaluations highlights its value in providing AI-assisted clinical decision support, where response accuracy, terminology precision, and contextual understanding are critical.
- **Descriptive Use Case:** AI-Driven Clinical Recommendations To assess its real-world effectiveness, GraphRAG is tested against complex clinical queries. When it is prompted with a query regarding the latest recommendations for managing insulin resistance in GDM patients, GraphRAG generates a structured response based on current guidelines and research findings.

The complete system response, which emphasizes evidence-based interventions and up-to-date clinical practices, is presented in the Box below.

#### GraphRAG Response to Query on Managing Insulin Resistance in GDM

***Query: What are the latest recommendations for managing insulin resistance in GDM patients?***

***GraphRAG Response: Recent studies suggest a multi-faceted approach to managing insulin resistance in GDM patients, including:***

**1.** Personalized low glycemic index diets, emphasizing complex carbohydrates and fiber-rich foods.
**2.** Structured exercise programs combining aerobic and resistance training for at least 150 minutes per week.
**3.** Early initiation of metformin where lifestyle modifications are insufficient, as per the 2023 guidelines.
**4.** Continuous glucose monitoring to optimize insulin dosing and adjust lifestyle recommendations.
**5.** Regular screening for comorbidities such as hypertension and dyslipidemia, which contribute to insulin resistance.

Unlike conventional LLMs, which tend to provide broad responses without citing specific guidelines, the GraphRAG local LLM integrates structured knowledge retrieval to generate responses that are both evidence-based and aligned with current clinical best practices. This level of detail ensures that healthcare professionals receive actionable insights rather than generic recommendations.

## 5. Conclusions and Future Work

The research reported in this paper demonstrates the promising potential of the GraphRAG framework as a clinical decision support tool for managing GDM. Trained on a curated dataset of published medical research, the GraphRAG-based local LLM significantly outperforms mainstream LLMs such as ChatGPT, Mistral, and Claude across multiple evaluation metrics. As evidenced in Figures 10 and 11, GraphRAG generates responses that are not only accurate and relevant but also contextually tailored to the specific demands of medical domains like GDM, especially within under-resourced settings in the Global South. Its superior BLEU, ROUGE, and Jaccard Similarity scores affirm its ability to replicate the linguistic precision and semantic fidelity required for clinical applications. In particular, the contributions of a GraphRAG-enabled local LLM are:

**1.** **Context-Aware Medical Content Generation:** It produces medically accurate content aligned with clinical guidelines, verified by strong performance in BLEU and ROUGE evaluations.
**2.** **Expert-Level Relevance and Precision:** The model synthesizes complex medical knowledge into clinically actionable responses, validated by expert reviewers for clarity and clinical reliability.
**3.** **Robust Handling of Complex Queries:** GraphRAG effectively addresses intricate topics such as insulin resistance and GDM pathophysiology, aiding informed clinical decisions.
**4.** **Scalable Clinical Support:** It maintains consistent response quality across diverse medical queries, fostering trust and usability in real-world clinical scenarios.

Beyond its technical performance, the approach introduces a novel integration of graph-based retrieval with local LLMs to support decentralized healthcare systems. This architecture allows healthcare providers—particularly general practitioners, nurse practitioners, and frontline clinicians—to access up-to-date medical insights without requiring deep specialization or an exhaustive literature review. Such functionality is critical in both high-demand clinical environments and underserved regions, where medical expertise is limited but decision-making remains time-sensitive. The capacity of GraphRAG local LLMs to bridge the informational gap between specialists and generalists highlight the transformative role in delivering equitable, scalable, and context-aware digital health support.

The demonstrated capabilities of GraphRAG in GDM management serve as a foundation for broader applications in medical AI. As we look to the future, such systems must be developed not to replace, but to complement human clinical judgment. We echo the caution raised in recent evaluations, such as (Liu et al. 2021), that equating LLM performance with physician competence through tests akin to the Turing Test is misguided. Medical AI—particularly generative models—must be architected to support clinicians, not supplant them. As Nambiar et al. (2024) saliently noted, the next phase lies in evaluating such tools prospectively as co-pilots in therapeutic decision-making, especially within fast-paced clinical environments. Two challenges are emphasized.

**1.** **Technical Innovations Contributing to Performance Gains**: GraphRAG’s superior performance in GDM-related decision support can be attributed to three primary technical innovations. The integration of KGs enhances response accuracy by mapping relationships between risk factors, symptoms, and treatment options. The RAG mechanism enables dynamic access to updated research, reducing the likelihood of outdated or incorrect recommendations. Also, domain-specific training allows GraphRAG to surpass general-purpose models in contextual understanding, ensuring that medical terminology and treatment pathways are accurately represented.
**2.** **Challenges and Considerations for Real-World Deployment**: Despite its promising results, integrating GraphRAG into clinical workflows presents several challenges. While the model shows strong alignment with medical knowledge, its adoption in real-world healthcare settings requires extensive validation through randomized controlled trials to ensure efficacy and reliability. Data privacy concerns must also be addressed, particularly in environments where sensitive patient data is involved. Interpretability remains another critical factor, as AI-generated recommendations must be transparent and explainable to gain trust among healthcare professionals. Moreover, the model’s ability to integrate with electronic health record systems and existing clinical decision support frameworks needs further exploration. The findings suggest that retrieval-augmented generative AI models such as GraphRAG have the potential to revolutionize AI-assisted healthcare decision-making. By bridging the gap between raw AI-generated text and structured clinical knowledge, such models pave the way for more reliable, evidence-based medical AI systems.

Future research should explore the scalability of this approach across other medical conditions, refining model architectures to improve generalization while maintaining domain-specific precision. The evaluation of GraphRAG highlights its potential as a specialized AI tool for GDM management, offering clinically relevant, research-backed recommendations with enhanced accuracy and contextual awareness. Its ability to synthesize structured knowledge retrieval with generative AI sets a new standard for AI-driven clinical decision support. However, further research is required to address implementation challenges, ensuring that such models can be effectively deployed in real-world healthcare environments. The integration of GraphRAG-like systems into mainstream medical practice could significantly enhance access to up-to-date medical knowledge, supporting more informed clinical decision-making and ultimately improving patient outcomes.

Future research should also focus on enhancing GraphRAG’s capabilities and expanding its applicability to a broader range of medical conditions. While the proof of concept demonstrates strong performance in GDM, deploying the model in real-world clinical settings presents several practical challenges^2^. These include computational overhead, the demand for high-quality domain-specific training data, and concerns related to model interpretability. Addressing these limitations is crucial to ensure seamless integration into clinical workflows and to support safe, explainable AI deployment in healthcare.

To improve the system’s retrieval precision and overall reliability, future work could integrate several technical advancements that may enhance GraphRAG’s real-world effectiveness. These include integrating advanced contextual retrieval methods, such as Contextual BM25 and embedding-based summarization, to ensure that retrieved data is both relevant and clinically significant. Additionally, retrieval-specific evaluation metrics such as failure rates and retrieval precision, should be integrated to expose gaps in the information pipeline. A more robust assessment of the retrieval process will provide valuable insights into the downstream quality of generative responses. Integrating re-ranking mechanisms to prioritize high-value clinical information, and leveraging hybrid retrieval strategies that combine semantic embeddings with lexical matching (e.g., BM25), may also improve performance in handling domain-specific medical terminology.

In parallel, legal and ethical considerations surrounding LLMs must be addressed^3^. One pressing concern is the potential for intellectual property (IP) infringement, particularly when using LLMs trained on proprietary or non-transparent datasets. To mitigate such risks, future iterations of GraphRAG will explicitly label and rely on open-source, publicly accessible datasets such as PubMed, in line with US and EU open-access mandates. This approach ensures transparency, reproducibility, and community trust.

Scalability and adoption in resource-constrained environments also require attention. Future work should explore strategies to reduce cost and latency through techniques such as prompt caching and efficient chunk management, ensuring system responsiveness without compromising accuracy or performance.

Privacy and data protection represent another critical axis of future development, particularly given recent debates over AI training on sensitive academic or patient data (Novelli, 2024). Incorporating privacy-preserving learning paradigms—such as federated learning and anonymized blockchains—can allow decentralized training without compromising confidentiality. Federated learning enables model improvement across multiple institutions without direct data sharing, helping to comply with regulations like the General Data Protection Regulation (GDPR) while protecting both patient identity and institutional data rights.

Equally important is the mitigation of model bias and hallucinations, especially in sensitive medical contexts. Techniques such as external data validation, selective re-ranking of outputs, and the use of diverse, high-quality datasets will help ensure the consistency, reliability, and equity of clinical guidance. By combining federated learning with strategies like off-chaining personally identifiable information, GraphRAG can be scaled across heterogeneous medical datasets while preserving data integrity and trust.

Looking furtherin to the future, GraphRAG may be extended globally to support patient-centric and personalized medicine. Integrating the system with real-time patient records in clinical workflows could enable dynamic, context-aware decision support systems that improve diagnostic accuracy and treatment planning. Following models such as AIDA, it would be valuable to evaluate GraphRAG prospectively as a clinical co-pilot in diagnosis and therapy, rather than as a replacement for human decision-making. Customizing GraphRAG outputs to reflect individual patient profiles—including genetic, environmental, and lifestyle factors— offers a pathway toward personalized medical guidance. These developments could meaningfully advance precision medicine and optimize healthcare delivery across diverse populations.

| **A Generational Account of Gestational Diabetes** |
|---|
| *“I do not wish to alarm you, Mrs. Sharma, but you have been diagnosed with gestational diabetes and your baby is 10 pounds at birth. Both of you need to be careful.”* |
| *— Matron in Singapore’s Kandang Kerbau Maternity Hospital to the mother of the last author, circa 1961.* |
| In 2022, the mother passed away peacefully at the age of 88, her diabetes controlled with insulin injections. The “baby” (the last author and principal investigator of this study) was diagnosed with type 2 diabetes at age 60, giving rise to the question of whether a GraphRAG solution may have changed the outcome for both with timely, relevant best practices. |

We conclude with the above vignette of gestational diabetes which serves as a poignant reminder of the enduring and often underestimated consequences of delayed or suboptimal maternal care. It reinforces our original question: *can early, accurate diagnosis and targeted intervention for GDM help prevent chronic disease progression and improve long-term outcomes, particularly in underserved communities?* This research suggest that AI solutions may indeed hold the key to addressing these gaps in regions where specialist expertise is scarce—such as in parts of the Global South. GraphRAG local LLMs’ context-aware, literature-informed support offer a practical means of augmenting clinical capacity. Even in resource-rich settings, physicians are increasingly burdened by the velocity of emerging medical knowledge. An AI Assistant like the PoC developed in this paper, designed to keep pace with evolving literature while maintaining clinical relevance, could provide timely, reliable guidance in everyday practice. To close, the evolution of contextual, graph-augmented language models offer a pathway to more adaptive, equitable, and scalable healthcare systems. Through responsible development and rigorous validation, these models can enhance diagnostic accuracy, reduce information asymmetries, and ultimately improve patient care—across generations. There is no question that the treatment of T2D and nested within, GDM, are in critical need of new approaches utilizing AI, digital technologies and valid datasets.

## Data Availability

All data produced are available online at https://github.com/fathima-ruba/GraphRAG-Architecture-of-a-local-LLM-for-Gestational-Diabetes-Mellitus

## Acknowledgments

This research was supported by the Research Incentive Fund (RIF) Grant R22086 from Zayed University to AN, RS & EE. The authors are grateful to the three medical practitioners—Dr. Z (a diabetologist), Dr. S (an ER physician), and Dr. R (a general physician)—for their thoughtful contribution to the prompts and looking over the GraphRAG responses for face validation. Many thanks are also due to the anonymous reviewers for guiding the revisions of the manuscript. AN and RS conceived the research idea. FR and SB conducted the empirical data collection and validation that produced the GraphRAG model. EE supervised the research and subsequent reporting as co-PI. All authors contributed equivalently to the research, analysis, and writing of this article. No human subjects nor live patient data were used to develop the PoC.

The data files for the project may be found at the following reference: F. Ruba, A. Nazir, E. Evangelista, S. Bukhari, L. bin Mohd Loft & R. Sharma. (2025.) Data Repository for GraphRAG-Architecture-of-a-local-LLM-for-Gestational-Diabetes-Mellitus. Available at: https://github.com/fathima-ruba/GraphRAG-Architecture-of-a-local-LLM-for-Gestational-Diabetes-Mellitus

## Disclosure statement

No potential conflict of interest is reported by the authors

## Footnotes

1 Gupta, A. (2025, April 22). AI could cure all disease in a decade, says Google DeepMind CEO— Perplexity’s Aravind Srinivas agrees. Mint. https://www.livemint.com/technology/tech-news/ai-could-cure-all-disease-in-a-decade-says-google-deepmind-ceo-perplexity-s-aravind-srinivas-agrees-11745285669206.html

2 Sam Altman Says AI Using Too Much Energy, Will Require Breakthrough Energy Source. (17-Jan-24). Futurism. https://futurism.com/sam-altman-energy-breakthrough.

3 Frermann, L., & Shaanan Cohney. (4-Feb-25). OpenAI says DeepSeek “inappropriately” copied ChatGPT – but it’s facing copyright claims too. The Conversation. https://theconversation.com/openai-says-deepseek-inappropriately-copied-chatgpt-but-its-facing-copyright-claims-too-248863.

## Notes

### Competing Interest Statement

The authors have declared no competing interest.

